# Rapid, widespread, and preferential increase of SARS-CoV-2 B.1.1.7 variant in Houston, TX, revealed by 8,857 genome sequences

**DOI:** 10.1101/2021.03.16.21253753

**Authors:** James M. Musser, Randall J. Olsen, Paul A. Christensen, S. Wesley Long, Sishir Subedi, James J. Davis, Jimmy Gollihar

## Abstract

Genetic variants of the SARS-CoV-2 virus have become of great interest worldwide because they have the potential to detrimentally alter the course of the SARS-CoV-2 pandemic, and disease in individual patients. We recently sequenced 20,453 SARS-CoV-2 genomes from patients with COVID-19 disease in metropolitan Houston (population 7 million), dating from March 2020 to early February 2021. We discovered that all major variants of concern or interest are circulating in the region. To follow up on this discovery, we analyzed 8,857 genome sequences from patients in eight Houston Methodist hospitals dispersed throughout the metroplex diagnosed from January 1, 2021 to March 7, 2021. This sample represents 94% of Houston Methodist cases and 4.8% of all reported cases in metropolitan Houston during this period. We discovered rapid, widespread, and preferential increase of the SARS-CoV-2 UK B.1.1.7 throughout the region. The estimated case doubling time in the Houston area is 6.9 days. None of the 648 UK B.1.1.7 samples identified had the E484K change in spike protein that can cause decreased recognition by antibodies.

## Introduction

Genetic variants of the SARS-CoV-2 virus are now of considerable concern internationally because some have been reported to spread rapidly, cause unusually severe infections, and be less controlled by naturally acquired or vaccine-induced immunity.^1-6^ By sequencing 20,453 SARS-CoV-2 genomes dating from March 2020 to early February 2021, we recently demonstrated all major variants of concern (VOCs) or interest (VOIs) are circulating in metropolitan Houston (population 7 million).^4^ These variants emerged in Houston in late December or early January. In the aggregate, the variants constituted only 1.1% of strains causing COVID-19 disease among patients in the eight Houston Methodist hospitals geographically dispersed throughout the metropolitan region. We identified 230 patients infected with a VOC or VOI, including 23 cases caused by B.1.1.7, a VOC reportedly increasing in frequency in several regions of the United States.^5^

Given their potential to detrimentally alter the course of the SARS-CoV-2 pandemic, and disease in individual patients, we tested the hypothesis that VOC and VOI are increasing in disease frequency in metropolitan Houston. We also sought to determine if any of the VOC/VOI caused an unusually severe clinical course.

## Methods

We sequenced the genomes of 8,857 SARS-CoV-2 from patient samples acquired January 1, 2021 through March 7, 2021 (**Figure 1A**) using Illumina instruments.^4^ This sample represents 94% of the 9,445 cases of COVID-19 diagnosed in the Houston Methodist system during this period. VOCs were identified based on genome sequence data and designated by Pangolin (https://cov-lineages.org/pangolin.html). Patient metadata were acquired from the electronic medical record by standard informatics methods. Patient home address zip codes were used to visualize the geospatial distribution of the B.1.1.7 VOC. Figures were generated with Tableau version 2020.3.4 (https://www.tableau.com/).

**Figure 1.**
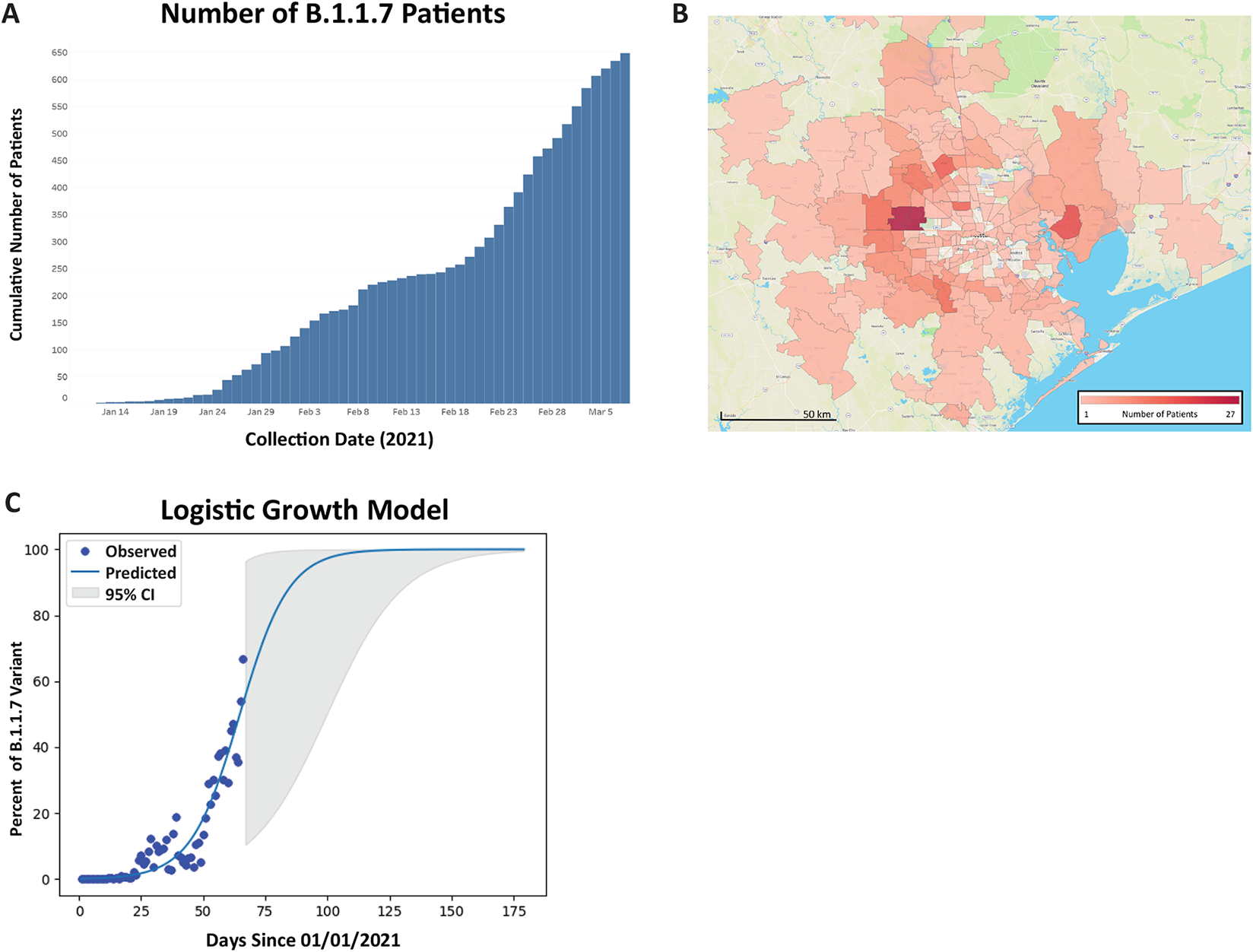
Epidemiologic curve, geospatial distribution, and growth model for the UK B.1.1.7 VOC in metropolitan Houston, Texas. **A**, Epidemiologic curve of COVID- 19 cases in the Houston Methodist hospital system. The curve shows the increased frequency of cases caused by B.1.1.7 from January 1, 2021 to March 7, 2021. **B**, Geospatial distribution of B.1.1.7 cases identified in the study. The home address zip code for each patient was used and figures were generated using Tableau version 2020.3.4. **C**, Growth model for the B.1.1.7 variant in metropolitan Houston. The relative growth of the B.1.1.7 variant was fitted to logistic growth model using the curve fit procedure with nonlinear least-squares estimation in SciPy (1.1.0) (https://docs.scipy.org/doc/scipy/reference/release.1.1.0.html). Growth rate: 0.1012 [95% confidence interval (CI): 0.0631 - 0.1393]; doubling time: 6.91 days (95% CI: 5.02 - 11.08).

## Results

We identified 648 cases caused by VOC B.1.1.7 and discovered that the variant increased in frequency during the study period (**Figure 1A**); in contrast, the other VOCs/VOIs did not (data not shown). Patients infected with B.1.1.7 were distributed throughout the entire metropolitan region (**Figure 1B**). Together, these data indicate rapid, widespread, and preferential dissemination of B.1.1.7 in the Houston area, with an estimated case doubling time of 6.9 days (**Figure 1C**). None of the 648 B.1.1.7 VOC had the E484K change in spike protein that can cause decreased recognition by antibodies.

To gain additional insight into these B.1.1.7 cases, we examined available patient metadata. Compared to non-B.1.1.7 cases, there was no significant difference in patient age, gender, median length of stay, or mortality (**Table 1**). However, significantly more patients with B.1.1.7 were hospitalized. Nasopharyngeal specimens from patients with the B.1.1.7 VOC had significantly lower Ct values (frequently used as a proxy for virus load) compared to non-B.1.1.7 cases, suggesting higher virus loads on initial diagnosis (**Table 1**). This finding is consistent with increased transmissibility of B.1.1.7.

**Table 1.**
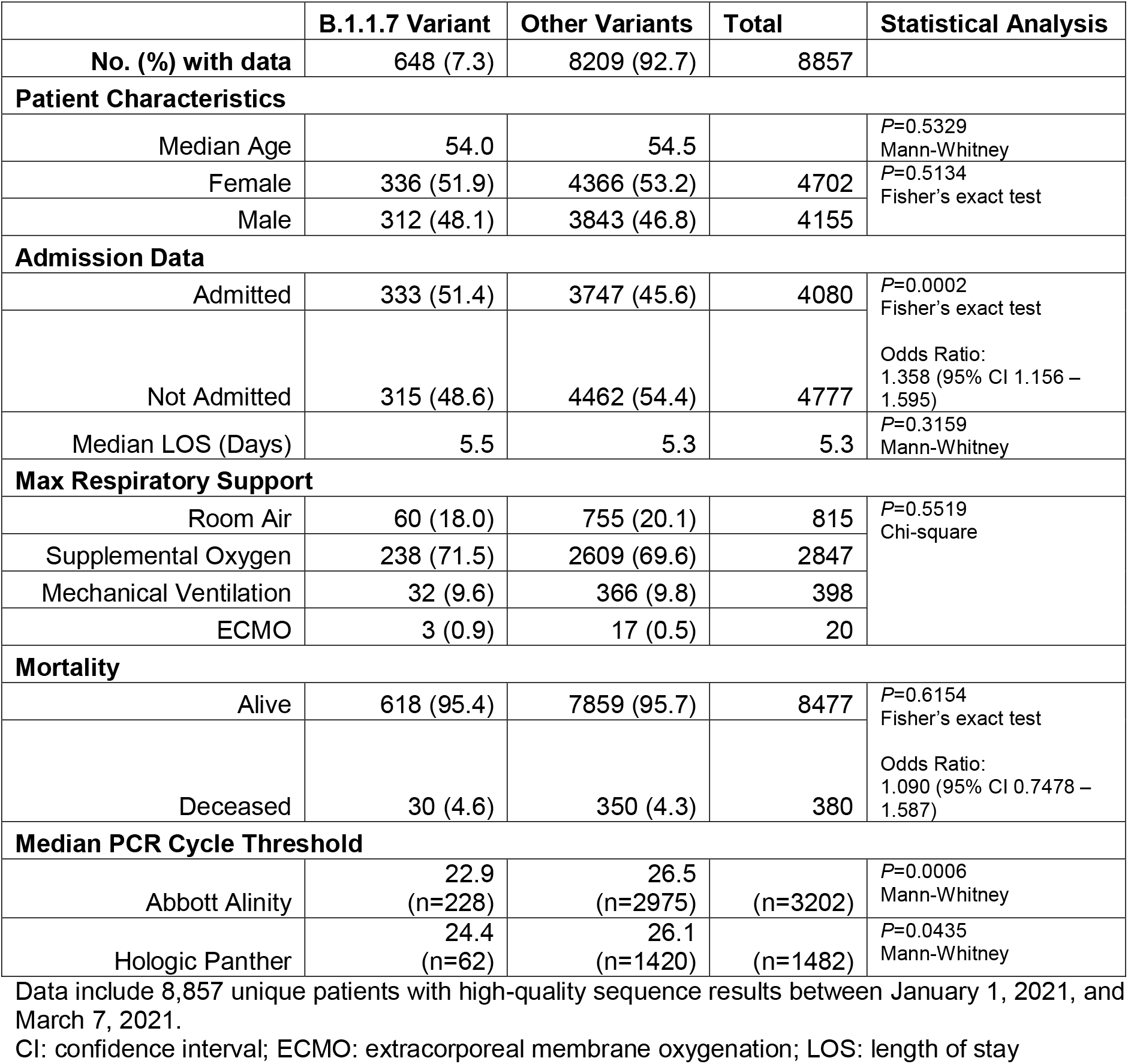
Summary of pertinent patient metadata for the 8,857 unique patients.

## Discussion

Our genome sequence data document a very rapid upward trajectory of VOC B.1.1.7 cases in metropolitan Houston. Importantly, there was no analogous increase in other variants. Our study has several limitations. During the study period, 197,817 cases of COVID-19 were reported in Harris County and eight contiguous counties (https://usafactsstatic.blob.core.windows.net/public/data/covid-19/covid_confirmed_usafacts.csv; last accessed March 12, 2021). Thus, our genome sample represents 4.8% of all cases in the metropolitan region and 94% of Houston Methodist cases. Our eight hospitals are widely distributed across the metropolitan region and serve patients who are demographically and geographically highly diverse, but it is likely our sample underrepresents SARS-CoV-2 genotypes circulating in difficult-to-reach populations, such as homeless and otherwise disenfranchised individuals. Finally, our hospitals care mainly for adult patients, which means that SARS-CoV-2 variants infecting pediatric patients are underrepresented in our study, although these are rare.

## Data Availability

Genome data used in this study have been deposited to GISAID.

## Author Contributions

Dr. Musser had full access to all study data and takes responsibility for the integrity of the data and the accuracy of the data analysis. *Concept and design:* Musser, Olsen, Christensen, Long

*Data acquisition, analysis, or interpretation of data*: All authors

*Drafting of the manuscript:* All authors

*Statistical analysis*: Christensen

*Obtained funding:* Musser and Davis

*Supervision:* Musser

*Authorship*: Musser, Olsen, Christensen, and Long contributed equally and are co-first authors

## Institutional Review Board statement

This work was approved by the Houston Methodist Research institutional review board (IRB1010-0199).

## Disclaimer

The findings and conclusions in this article are those of the authors and do not necessarily reflect the views of the U.S. Army.

## Additional Contributions

We are indebted to Drs. Marc Boom and Dirk Sostman for their ongoing support. We thank Matthew Ojeda Saavedra, Prasanti Yerramilli, Layne Pruitt, Kristina Reppond, Madison N. Shyer, Jessica Cambric. Dr. Ilya J. Finkelstein, Dr. Jessica W. Podnar, Hung-Che Kuo and G. Nguyen for genome sequencing support. We acknowledge the assistance of Robert Olson, Maulik Shukla and Marcus Nguyen at the Bacterial and Viral Bioinformatics Resource Center for their assistance with sequence analysis. Dr. Parsa Hodjat contributed to formulating the real-time Tableau dashboard analysis. We thank Drs. Kathryn Stockbauer and Sasha Pejerrey for editorial contributions.

## Additional Information

Genome data used in this study have been deposited to GISAID.

